## Supplementary material for "Design of the HPV-Automated Visual Evaluation (PAVE) Study: Validating a Novel Cervical Screening Strategy": PAVE Committees

| **Date: September 24th 2023** | **Brief summary** |
| --- | --- |
| **Project Leads:** Mark Schiffman and Silvia de Sanjosé | **Scientific leadership. To overview the project, guarantee ethical consideration, traceability of the steps, and accountability. Response to the funders, and planning for results dissemination.** |
| **Coordination and site overview**: Mark Schiffman, Silvia de Sanjose, Kanan Desai and Federica Inturrisi | **Nigeria: Mark Schiffman & Kanan Desai**  **Brazil: Mark Schiffman & Anna Cecilia Rodriguez**  **Tanzania, Malawi & Eswatini: Federica Inturrisi**  **El Salvador, Honduras, Dominica Republic and Cambodia: Silvia de Sanjosé** |
| **Epidemiology and Biostatistics**:  **Ana Cecilia Rodriguez**, **Didem Egemen**, Brian Befano,  Li Cheung, Nicole Campos | **Planning and running the statistical analyses of PAVE strategy** |
| **AI**: **Jayashree Kalpathy-Cramer**,  Rakin S. Ahmed, Brian Befano, Didem Egemen, Ana Cecilia Rodriguez | **Evaluation of algorithm performance for quality, diagnosis and SCJ identification** |
| **Data coordination (IMS)**: **Jen Boyd-Morin**, **Brian Befano** | **Data final reception and data analysis. Image review for quality and delivery for AI evaluation.** |
| **Clinical, Pathology, Treatability and QA**: **Rebecca Perkins**, **Akiva Novetsky**, Diego Guillén, Jose Jerónimo,  Jenna Marcus, Kanan Desai, Ana Cecilia Rodriguez | **Assessment of the adequacy of clinical protocol, pathology external review and follow up of patients in need for treatment. It includes the endpoint adjudication** |
| **Data Collection and Field Integration**: **Silvia de Sanjosé**, Kanan Desai, Ana-Cecilia Rodriguez, EHAS | **On site data and image collection using a DHIS2 app designed for PAVE or comparable apps providing a pre-specified data set.**  **EHAS (Ignacio Prieto leads the coordination in all PAVE sites, checks the data for completeness and transfers the data to IMS)** |
| **HPV**: **Mark Schiffman**, Kanan Desai, Taina Raiol , Federica Inturrisi | Close monitoring of the ScreenFire performance in each site. Carry QC studies to guaranty high accuracy |
| **Cost-Effectiveness**: **Nicole Campos** and site-specific collaborators | **Micro-costing in four study sites including cost to train** |
| **HIV: Silvia de Sanjosé,** José Jerónimo, Federica Inturrisi and collaborators | **To guarantee that WLWH are clearly represented and evaluated within PAVE** |
| **COVID Risk Mitigation**: **Didem Egemen**, Silvia de Sanjosé | **Providing instructions/ recommendation to avoid COVID transmission** |
| **Communication and Retention: Paul Han,** Abigail Ukwuani, Imran Mohhason- Bello, Zeev Rosberger, Natasha Hansen, Karen Yates, Montserrat Garcia | **Evaluating knowledge, risk perceptions at the provider, stakeholders and user levels.** |
| **Steering Committees:**  **All PIs in each site and leaders of the Committees** |  |

| **Date:26 sept 2022** | **Brief summary** |
| --- | --- |
| **Project Leads:** Mark Schiffman and Silvia de Sanjosé | The Project is moving along, working on contracts ScreenFire, Copan, IRIS |
| **Epidemiology and Biostatistics**:  **Ana Cecilia Rodriguez**, **Didem Egemen**, Brian Befano, Julia Gage, Li Cheung, Nicole Campos | NTR |
| **AI**: **Jayashree Kalpathy-Cramer**,  Rakin S. Ahmed, Brian Befano, Didem Egemen, Ana Cecilia Rodriguez | Papers moving along. Unknown outcome of the Zambia paper |
| **Data coordination (IMS)**: **Jen Boyd-Morin**, **Brian Befano** | **NTR** |
| **Clinical, Pathology, Treatability and QA**: **Rebecca Perkins**, **Akiva Novetsky**, Diego Guillén, Jose Jerónimo,  Jenna Marcus, Kanan Desai, Ana Cecilia Rodriguez | Quality algorithm moving along; Treatability algorithm is advancing |
| **Data Collection and Field Integration**: **Silvia de Sanjosé**, Kanan Desai, Ana-Cecilia Rodriguez, EHAS | **NTR** |
| **HPV**: **Mark Schiffman**, Kanan Desai, Taina Raiol , Federica Inturrisi | Experiments on ScreenFire proceedings are underway in Frederick |
| **Cost-Effectiveness**: **Nicole Campos** and site-specific collaborators | The training course is ongoing with good attendance. |
| **HIV, FGS**: **Helen Kelly**, José Jerónimo, Federica Inturrisi and collaborators | Brief summary of the activities with john D. Maybe interesting to present an overall summary of the results |
| **COVID Risk Mitigation**: **Didem Egemen**, Silvia de Sanjosé | **NTR** |
| **Communication and Retention: Paul Han,** Abigail Ukwuani, Imran Mohhason- Bello, Zeev Rosberger, Natasha Hansen, Karen Yates, Montserrat Garcia | Proposal to start questionnaire on risk communication with the PAVE researchers |
| **Coordination**: **Farideh Almani**, | Update on the need to request clearance once a paper is ready fro submission |
| **Other topics: all** | NTR |
