## Supplementary material on Impact of Vaccination adn Screnning and on data sharing process for "Design of the HPV-Automated Visual Evaluation (PAVE) Study: Validating a Novel Cervical Screening Strategy"

**Supplementary Materials to**

**“HPV-Automated Visual Evaluation (PAVE) Study: Design and methods”**

**Supplementary Methods: Projecting the cost and health impact of cervical screening and single-dose HPV vaccination in the highest burden countries.**

Our objective was to project the relative timing of health benefits and the cost per cervical cancer death averted following cervical screening with HPV-based screening and prophylactic single-dose HPV vaccination of adolescents in low-resource countries with a high burden of cervical cancer. Two scenarios were examined: 1) a one-time highly effective screening campaign of 30- 49 year-old women in 2027 (i.e., 20 birth cohorts); and 2) vaccinating 90% of 9- 14 year-old girls in 2027 (i.e., 6 birth cohorts) with a bivalent HPV16/18 vaccination. We selected all low- and lower-middle income countries with an age-standardized cervical cancer incidence rate greater than 10 per 100,000 women (Supplemental Table S1; n=67). For each country, we assumed that, in the absence of any intervention, the number of cervical cancer deaths for each 5-year age group would apply each year for the lifetime of the selected birth cohorts.^1^ We conservatively and crudely assumed that screening and management would avert 25% of cervical cancer deaths (equivalent to screening uptake of 40% of eligible women, with 62.5% of screen-positive women receiving appropriate management) beginning at age 50 years. For vaccination cohorts, we assumed that a bivalent HPV16/18 vaccine (i.e., against the genotypes responsible for 70% of cervical cancers) with 90% uptake would avert 63% of cervical cancer deaths.

While data on the costs of implementing novel screening strategies and single-dose HPV vaccination for female adolescents are forthcoming from the PAVE consortium and single-dose vaccination studies, we crudely assumed a single vaccine dose cost US$4.50^2^ with an average financial delivery cost per dose (i.e., per fully immunized girl) of US$7.^3^ We assumed a bundled financial cost per woman screened of US$15, including a low-cost rapid HPV genotyping assay with triage and treatment of screen-positive women.

According to our projections, the number of interventions needed to avert one cervical cancer death was similar for HPV vaccination and screening (i.e., 278 for HPV vaccination; 293 for screening). A onetime screening campaign for women aged 30 to 49 years in the selected countries yielded a financial cost of ~US$2.5 billion to avert ~570,000 deaths, or US$4,400 per death averted. On a similar order of magnitude, a onetime single-dose bivalent HPV vaccination campaign of girls aged 9 to 14 years in the same countries would cost ~US$2.0 billion and avert ~640,000 deaths, or US$3,200 per death averted. Of note, these ballpark estimates are undiscounted and do not account for cancer treatment cost offsets. We also did not consider demographic changes over the lifetime of intervention cohorts, nor did we consider indirect benefits of vaccination or prevention of other HPV-related cancers.

**Supplementary References**

**Supplementary Table S1. Selected low- and lower-middle income countries with age-standardized cervical cancer incidence rates >10 per 100,000 women (ranked by decreasing age-standardized incidence).**

| Country | World Bank Income Group^4^ | Age-standardized cervical cancer incidence rate per 100,000 women^1^ |
| --- | --- | --- |
| Eswatini | LMI | 84.5 |
| Malawi | LI | 67.9 |
| Zambia | LI | 65.5 |
| Tanzania | LMI | 62.5 |
| Zimbabwe | LMI | 61.7 |
| Lesotho | LMI | 56.8 |
| Uganda | LI | 56.2 |
| Comoros | LMI | 56.0 |
| Mozambique | LI | 50.2 |
| Guinea | LI | 50.1 |
| Burundi | LI | 49.3 |
| The Republic of the Gambia | LI | 42.9 |
| Madagascar | LI | 41.2 |
| Liberia | LI | 40.8 |
| Guinea-Bissau | LI | 39.6 |
| Angola | LMI | 37.6 |
| Bolivia (Plurinational State of) | LMI | 36.6 |
| Mali | LI | 36.4 |
| Cameroon | LMI | 33.7 |
| Congo, Democratic Republic of | LI | 31.9 |
| Kenya | LMI | 31.3 |
| Côte d’Ivoire | LMI | 31.2 |
| Papua New Guinea | LMI | 29.2 |
| Mauritania | LMI | 28.9 |
| Rwanda | LI | 28.2 |
| Ghana | LMI | 27.4 |
| Solomon Islands | LMI | 25.4 |
| Somalia | LI | 25.1 |
| Indonesia | LMI | 24.4 |
| Myanmar | LMI | 22.6 |
| Congo, Republic of | LMI | 22.4 |
| Central African Republic | LI | 21.8 |
| Ethiopia | LI | 21.5 |
| Nicaragua | LMI | 21.3 |
| Sierra Leone | LI | 21.2 |
| South Sudan | LI | 20.5 |
| Chad | LI | 20.2 |
| Mongolia | LMI | 19.7 |
| Honduras | LMI | 19.5 |
| Togo | LI | 19.1 |
| Nigeria | LMI | 18.4 |
| Burkina Faso | LI | 18.2 |
| India | LMI | 18.0 |
| Vanuatu | LMI | 17.1 |
| Cabo Verde | LMI | 17.0 |
| Nepal | LMI | 16.4 |
| Kyrgyzstan | LMI | 15.4 |
| Djibouti | LMI | 15.3 |
| Eritrea | LI | 15.3 |
| Philippines | LMI | 15.2 |
| Benin | LMI | 15.1 |
| Ukraine | LMI | 14.3 |
| Bhutan | LMI | 14.2 |
| Cambodia | LMI | 14.0 |
| Timor-Leste | LMI | 14.0 |
| Micronesia/Polynesia | LMI | 16.3 |
| El Salvador | LMI | 131 |
| Samoa | LMI | 12.4 |
| Lao People’s Democratic Republic | LMI | 12.0 |
| Haiti | LMI | 11.6 |
| Korea, Democratic Republic of | LI | 11.2 |
| Uzbekistan | LMI | 11.0 |
| Bangladesh | LMI | 10.6 |
| Afghanistan | LI | 10. |
| Morocco | LMI | 10.4 |
| Niger | LI | 10.4 |

Annex

**Flow chart of data sharing in the PAVE study sites using the DHIS2 during the efficacy phase**

The figure bellow (Supplementary Figure S1) illustrates the various steps involved in the PAVE consortium, from data collection to data analysis. This outline is for those sites utilizing the District Health Information System 2 (DHIS2) app, where pre-defined data is collected from women who agree to participate. It is important to note that any individual information collected for clinical management purposes will not be uploaded to the study server

The study server incorporates the ITEA app, which facilitates the transfer of data to the NGO EHAS. At EHAS, the data undergoes a thorough check to ensure appropriate study identification. Additionally, if necessary, cervical images of the participants are linked to the core database. Once the verification process is approved, the data is sent to National Cancer Institute- Information Management System (NCI-IMS) for preparation before analysis.

During this step, data verification and cleaning procedures are performed to ensure the accuracy and quality of the data. Subsequently, the images, as well as the HPV and histology assessments (when available), are forwarded to Prof Jayashree Kalpathy-Cramer's team for AI evaluation. The results, in the form of data scores, are then transmitted back to NCI-IMS for comprehensive analysis of the consortium data.

Note: Sites using other apps like RedCap will extract the Core Data from their files and forward it to EHAS. Images will follow the same procedure.

Supplementary Figure S1. Flowchart of data sharing in the PAVE study sites

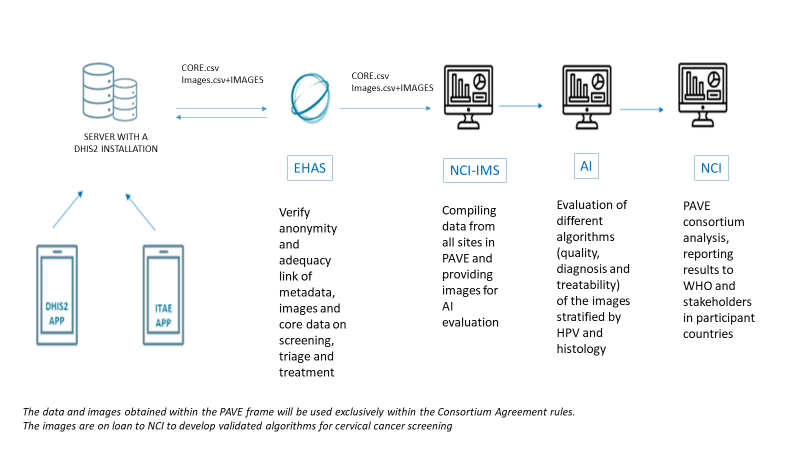

Appendix Roles and responsibilities

Names, affiliations, and roles of protocol contributors

See page 1

Name and contact information for the trial sponsor

Mark Schiffman, NCI

Role of study sponsor and funders, if any, in study design; collection, management, analysis, and interpretation of data; writing of the report; and the decision to submit the report for publication, including whether they will have ultimate authority over any of these activities

Composition, roles, and responsibilities of the coordinating centre, steering committee, endpoint adjudication committee, data management team, and other individuals or groups overseeing the trial, if applicable (see Item 21a for data monitoring committee)

Appendix Data collected

| **Column csv name** | **Form name** |
| --- | --- |
| NCI_PID | NCI PID |
| orgUnitName | Organization unit |
| Enrollment.date | Enrollment date |
| Enrollment.dataValues.CS (e) - Age | Age |
| Enrollment.dataValues.CS (e) - Number_vaginal_deliveries | Number of vaginal deliveries |
| Enrollment.dataValues.CS (e) - Last_menstrual_period_date | Last menstrual period |
| Enrollment.dataValues.CS (e) - HIV Status | HIV Status |
| Enrollment.dataValues.CS (e) - Signed_IC | Signed consent information |
| Enrollment.dataValues.CS (e) - HIV_Taking ART | Taking ART |
| HPV Screening-1.date | HPV Screening date |
| HPV Screening-1.dataValues.CS (h) - ID_test_HPV | HPV Test ID |
| HPV Screening-1.dataValues.CS (h) - Type_screening | Type of test |
| HPV Screening-1.dataValues.CS (h) - Specify_ other_test | Specify the other test |
| HPV Screening-1.dataValues.CS (h) - HPV_sampling | HPV Sampling |
| HPV Screening-1.dataValues.CS (h) - HPV_Result | Result |
| Triage-1.date | Triage date |
| Triage-1.dataValues.CS (tri)- VIA | VIA |
| Triage-1.dataValues.CS (tri)- VIA_result | VIA result |
| Triage-1.dataValues.CS (tri)- Other VIA result | Other |
| Triage-1.dataValues.CS (tri)- VAT | VAT |
| Triage-1.dataValues.CS (tri)- VAT_result | VAT Results - adequacy for ablative treatment |
| Triage-1.dataValues.CS (tri)- Cytology | Cytology |
| Triage-1.dataValues.CS (tri)- Cytology_result | Cytology result |
| Triage-1.dataValues.CS (tri)- Result date cytology | Result date |
| Triage-1.dataValues.CS (tri)- Colposcopy | Colposcopy |
| Triage-1.dataValues.CS (tri)- Colposcopy_result | Are visible lesion(s)? |
| Triage-1.dataValues.CS (tri)- Other_colposcopy_result | Other |
| Triage-1.dataValues.CS (tri) - Cervical_visualization_result | Cervix fully visualized |
| Triage-1.dataValues.CS (tri)- Cervical_visualisation_Bleeding | Bleeding |
| Triage-1.dataValues.CS (tri)- Cervical_visualisation _Ectopy | Ectopy |
| Triage-1.dataValues.CS (tri)- Cervical_visualisation_Mucus | Mucus |
| Triage-1.dataValues.CS (tri)- Cervical_visualisation_Prolapse | Uterine prolapse |
| Triage-1.dataValues.CS (tri)- Cervical_visualisation_Large | Large Cyst(s) |
| Triage-1.dataValues.CS (tri)- Cervical_visualisation_Strong | Inflammatory component |
| Triage-1.dataValues.CS (tri)- Cervical_visualisation_Infectious | Infectious visible component |
| Triage-1.dataValues.CS (tri) - SCJ_visualization | Squamocolumnar Junction (SCJ) visualization |
| Triage-1.dataValues.CS (tri)- Image capture | Image capture |
| Triage-1.dataValues.CS (tri)- Type of device - IRIS | IRIS |
| Triage-1.dataValues.CS (tri)- Type of device - Smartphone | Smartphone |
| Triage-1.dataValues.CS (tri) - Reason_no_image_capture | Reason for no images |
| Triage-1.dataValues.CS (tri)- Biopsy_taken | Were biopsies collected? |
| Triage-1.dataValues.CS (tri)- Reason no biopsy | Reason for no biopsies |
| Triage-1.dataValues.CS (tri)- Number_samples_biopsy_ectocervix | How many ectocervix samples? |
| Triage-1.dataValues.CS (tri)- Which_device_ectocer | Which device ectocervix? |
| Triage-1.dataValues.CS (tri)- Biopsies_date_result_ecto | Result Date Ectocervix |
| Triage-1.dataValues.CS (tri)- Result_biopsy-Normal | Normal |
| Triage-1.dataValues.CS (tri)- Result_biopsy-Cervicitis | Cervicitis |
| Triage-1.dataValues.CS (tri)- Result_biopsy-CIN1 | CIN1 |
| Triage-1.dataValues.CS (tri)- Result_biopsy-CIN2 | CIN2 |
| Triage-1.dataValues.CS (tri)- Result_biopsy-CIN3 | CIN3 |
| Triage-1.dataValues.CS (tri)- Result_biopsy-Carcinoma | Carcinoma |
| Triage-1.dataValues.CS (tri)- Result_biopsy-Adenocarcinoma | Adenocarcinoma |
| Triage-1.dataValues.CS (tri)- Result_biopsy-AIS | AIS |
| Triage-1.dataValues.CS (tri)- Result_biopsy-Other (specify) | Other(Specify) |
| Triage-1.dataValues.CS (tri) - Other_biopsy_ecto | Specify the other result |
| Triage-1.dataValues.CS (tri)- Result_biopsy-Sample insufficient | Sample insufficient |
| Triage-1.dataValues.CS (tri) - Clock First quadrant | First quadrant |
| Triage-1.dataValues.CS (tri) - Clock Second quadrant | Second quadrant |
| Triage-1.dataValues.CS (tri) - Clock Third quadrant | Third quadrant |
| Triage-1.dataValues.CS (tri) - Clock Fourth quadrant | Fourth quadrant |
| Triage-1.dataValues.CS (tri)- Sample_endocervix | Endocervical sample? |
| Triage-1.dataValues.CS (tri)- Which_device_endocer | Which device for endocervical? |
| Triage-1.dataValues.CS (tri)- Biopsies_date_result_endo | Result Date endocervical |
| Triage-1.dataValues.CS (tri) - Result_biopsy_endo | Result biopsy Endocervical |
| Triage-1.dataValues.CS (tri) - Other_biopsy_endo | Other Result biopsy endocervical |
| Triage-1.dataValues.CS (tri) - Management_triage | Management of the triage stage |
| Treatment-1.date | Treatment date |
| Treatment-1.dataValues.CS (tre)- Decision_treatment | Decision |
| Treatment-1.dataValues.CS (tre)- Number_applications | Number of applications |
| Treatment-1.dataValues.CS (tre)- Type_probe_16 mm | Type of probe - 16 mm |
| Treatment-1.dataValues.CS (tre)- Type_probe_19 mm | Type of probe - 19 mm |
| Treatment-1.dataValues.CS (tre)- Type_probe_19 mm_nipple | Type of probe - 19 mm nipple |
| Treatment-1.dataValues.CS (tre) - Other_treatment | Specify the other treatment |
| Treatment-1.dataValues.CS (tre) - Result_treatment | Treatment result |
| Treatment-1.dataValues.CS (tre)- Treatment_correctly_finished | Treatment correctly finished |
| Treatment-1.dataValues.CS (tre) - Reason_No_Correctly_Finished | Why wasn't the treatment correctly finished? |
| Follow up & End of study.date | Follow up and End of study |
| Follow up & End of study.dataValues.CS (f) - Finished | Case finished? |
| Follow up & End of study.dataValues.CS (f) - Missing_data | Missing data on: |
| Follow up & End of study.dataValues.CS (f) - Final_diagnosis | Final diagnosis |
| Adverse event.date |  |
| Adverse event.dataValues.CS (a) - Notified_coordinator_AE | Notified to the coordinator? |
| Adverse event.dataValues.CS (a) - Category_AE | Category |
| Adverse event.dataValues.CS (a) - Grade_AE | Grade |
| Adverse event.dataValues.CS (a) - Resolution_date_AE | Resolution date |
| RESULTS-1.S- ID HPV Test | HPV Test ID |
| RESULTS-1.date | Date |
| RESULTS-1.dataValues.SF (R) - HPV16-CY5 | HPV16-CY5 |
| RESULTS-1.dataValues.SF (R) - HPV18/45-ROX | HPV18/45-ROX |
| RESULTS-1.dataValues.SF (R) - HPV31/33/35/52/58-Vy5.5 | HPV31/33/35/52/58-Vy5.5 |
| RESULTS-1.dataValues.SF (R) - HPV39/51/56/59/68-FAM | HPV39/51/56/59/68-FAM |
| RESULTS-1.dataValues.SF (R) - Internal control-HEX | Internal control-HEX |
| RESULTS-1.dataValues.SF (R) - Date | Date |
